# The VIPR-1 trial (Visualizing Ischemia in the Pancreatic Remnant) - Assessing the role of intraoperative indocyanine green perfusion of the transected pancreas in predicting postoperative pancreatic leaks: protocol for a phase II clinical trial

**DOI:** 10.1101/2024.09.13.24313603

**Authors:** Gustavo Salgado-Garza, Annika Willy, Flavio G. Rocha, Skye C. Mayo, Brett C. Sheppard, Patrick J. Worth

## Abstract

Surgery of the pancreas has come a long way since its inception; however, postoperative morbidity is still high. Pancreatic leaks and fistulas are common complications in patients undergoing surgery to remove the pancreas. Fistulas delay subsequent oncological care after surgery and prolong the hospital stay. Hypoperfusion to the pancreas has been characterized as one factor leading to fistulas. Indocyanine green (ICG) injection allows the surgeon to evaluate blood perfusion to tissue in real-time. This protocol describes a trial that aims to assess the effectiveness of intraoperative ICG metrics of the cut edge of the pancreas to predict postoperative fistulas. A single group will participate in an observational, surgeon-blinded, phase II trial. ICG measurements of the cut edge of the pancreas will be recorded before reconstruction. International Study Group on Pancreatic Surgery criteria for pancreatic fistula will be used to define leaks and fistulas. The primary outcome will be the correlation between ICG measurements and the development or absence of fistula formation. Currently, limited objective intraoperative predictors exist for predicting postoperative fistulas. Having a reliable predictive tool could decrease the healthcare burden posed by fistulas. The findings of this trial will provide conclusions on the usefulness of ICG measurements in predicting postoperative pancreatic fistulas and leaks. This clinical trial is registered in ClinicalTrials.gov with the ID NCT06084013. The current protocol version is v1.0.

## Introduction

Pancreatic cancer is a deadly disease with a growing incidence rate, causing over 50,000 deaths annually in the United States [1]. The 5-year survival rate for this cancer is very low, but it can increase to over 30% with successful surgical resection [2]. However, one of the most serious complications of pancreatic surgery is the development of postoperative pancreatic fistulas (POPF) or leaks, which can occur in up to 1 in 3 patients [3]. The impact of POPF on patients is twofold; it not only increases immediate postoperative complications but also adversely affects medium to long-term oncological outcomes. POPF increases postoperative mortality because of prolonged hospital stays and higher reoperation rates [4]. In addition, POPF is also linked to worse oncological outcomes for patients, notably tumor recurrence and overall survival [5,6].

Current fistula risk prediction models do not include modifiable intraoperative risk factors for mitigating pancreatic fistula [7]. The poorly perfused surgical neck of the pancreas, vulnerable to ischemia, is a known key factor in developing leaks [8,9]. Indocyanine green (ICG) fluorescence is used to assess tissue perfusion during surgery, but its effectiveness in predicting pancreatic leaks has yet to be systematically studied. A reliable intraoperative method to analyze blood perfusion could help guide surgical margins and reduce the risk of pancreaticojejunostomy leaks, ultimately improving patient outcomes.

Indocyanine green (ICG) is an FDA-approved dye that emits fluorescence when near-infrared light is applied, making it useful for visualizing tissue perfusion during surgery. ICG is injected intravenously, traveling to tissues via arteries. While ICG is now considered standard in some surgical disciplines, it currently needs to be widely used in pancreatic surgery, primarily due to a lack of adequately powered prospective studies analyzing the benefits of its use and surgeons wanting more evidence to change their practices [10]. Regarding pancreatic surgery, ICG is mostly used for anatomical identification of structures to improve surgical safety, avoid damage to critical structures, and delineate tumors for excision [11–13].

In pancreatic surgery, ICG has been used to visualize tissue blood perfusion, with recent studies showing promising results [13,14]. Pancreatic surgeons have shown interest in further studies on ICG for better visualization of tissue perfusion. A recently published Delphi consensus study reported that almost 80% of experts agreed on the need to focus research efforts of ICG in predicting the risk for POPF [10]. Although research on ICG for evaluating leaks after pancreatectomy is limited, one study showed its potential for detecting leaks in a single patient retrospectively [13]. ICG has shown potential benefits in improving visualization and outcomes in pancreas surgery. Moreover, ICG has an excellent safety profile, with Clavien-Dindo grade ≥4 complications occurring in only 0.05% of cases where it is utilized [15]. Long-term potential benefits include correlating ICG uptake with pancreatic remnant perfusion and providing critical information for clinical decisions to avoid POPFs.

This study protocol aims to investigate the relationship between hypoperfusion during surgery and the development of postoperative leaks to identify modifiable intraoperative interventions to decrease leak rates. The goal is to leverage the potential positive outcomes from this study to design a phase III clinical trial to evaluate improving remnant gland perfusion and potentially decreasing complications.

## Materials and methods

This protocol is for a Phase II, open-label, surgeon-blinded, observational study to assess the predictive potential of intraoperative perfusion parameters at the pancreatic cut edge via ICG and its relation to fistula formation versus conventional measurements. The study design consists of a single-arm intervention group at a single academic center: the Oregon Health & Science University Hospital in the United States. Following international and national definitions, this center is considered a high-volume hospital for pancreatectomies [16,17].

This study protocol adheres to the SPIRIT guidelines for trial reporting [18]. To ensure adequate representation of ethnic and racial group demographics in the study, a full consent form and protocol will be available for English- and Spanish-speaking participants. Short summaries of the consent forms are available in other languages. Consent will be obtained by members of the surgical department, including clinical staff and research assistants. This clinical trial is registered in ClinicalTrials.gov with the ID NCT06084013.

### Sample size

Given the limited data of ICG perfusion in the pancreas, the sample size calculation was based on determining a detectable difference between the fistula and no fistula groups. Anticipating balanced numbers between the two groups, our sample size of 50 participants will have 80% power to detect at least a 25% difference in a leak or fistula rate, assuming a 20% leak rate for a normal ICG perfusion group. We account for a liberal exclusion of 30% of participants due to multiple and complex postoperative factors known to be associated with leaks. Therefore, we anticipate screening 75 participants and enrolling 50.

### Eligibility Criteria

All participants must fulfill eligibility criteria and have none of the excluding criteria. Inclusion Criteria:

1. Participant scheduled for pancreaticoduodenectomy for any diagnosis.
2. Participants≥ 18 years of age.
3. Ability to understand the nature and individual consequences of clinical trials.
4. Written informed consent from the participant or legally authorized representative.
5. For participants of childbearing potential, a negative pregnancy test and adequate contraception until 14 days after the trial intervention.
6. Participant needs to have an operative drain (any closed suction drain) after the procedure.
7. Participants that do not require arterial reconstruction.
8. Participants that require minor portal venous reconstruction including patch venoplasty.

### Exclusion Criteria

1. Patients with previous history of adverse reaction to contrast dye, ICG or components of the dye.
2. Prior pancreatectomy.
3. Known diagnosis of hepatic insufficiency, hepatitis, liver fibrosis or cirrhosis, or chronic pancreatitis.
4. Because this study focuses on hypoperfusion, patients will be excluded if in postoperative day 3-5 had any of the following: persistent SBP <90 mmHg unresponsive to 1L crystalloid, unexpected ICU transfer, blood transfusion of >2 units intraoperatively or 1 postoperatively, continuous vasopressor treatment or ACLS protocol initiation.
5. Organ failure, anuria or NSQIP-identified complication will be reviewed by PI and attending surgeon and decide exclusion.
6. Patients that require arterial reconstruction as part of their procedures.

### Blinding

To avoid the influence of ICG measurements on surgical decisions at this stage of the trial, this study will involve surgeon-blinding. During intraoperative measurements, the primary surgeon must either step outside the operating room or be positioned to avoid a line of sight to the imaging device monitor. The secondary surgeon will remain scrubbed in for this portion of the case. Both study personnel and operating room staff will ensure this blinding process is successful. Any instances of the surgeon viewing the measurements will result in the participant’s removal from the study. To ensure adequate surgeon-blinding, objective measurements of the captured images are performed postoperatively, with no relay of this information to the surgeons during the 30 days of the active study phase. Since this study is open-label, participants will know if they received the dye and underwent intraoperative measurements. As this intervention is not standard of care, there is no need for premature unblinding to the surgeons. Imaging measurements will only be available to surgeons and participants after the end of the study follow-up, which is 30 days after surgery.

### Intervention

During pancreatectomy, after the surgical specimen is removed and before creating the pancreatojejunostomy anastomosis, a single dose of 12.5 mg (5mL once reconstituted) of ICG will be administered intravenously by the anesthesiologist or certified nurse anesthetist. Spy+ Elite (Stryker, USA) will be used to record perfusion metrics, taking as reference the gastric body and small intestine to compare differences in perfusion of the pancreatic stump. Recording of a video image starts as soon as the dye is injected. Still pictures will be extracted from the video at 10-second intervals once tissue saturation is stable, allowing a comparison of perfusion to the pancreatic stump and the reference tissue.

While measurements and recording of perfusion are being obtained, the primary surgeon will step out of the operating room to ensure that ICG-derived data does not influence the surgical plan. Upon the study’s completion, descriptive statistics will be reported to analyze the impact of ICG use and perfusion measurements on pancreatic surgeries. During ICG administration, vital signs will be continuously monitored, and hypersensitivity symptoms will be managed promptly. An intra-abdominal drainage tube will be placed to assess for amylase levels.

Participants can withdraw consent at any time. Adverse effects from the study interventions will be monitored and reported to institutional authorities. Figure 1 shows the schedule for participants.

**Figure 1.**
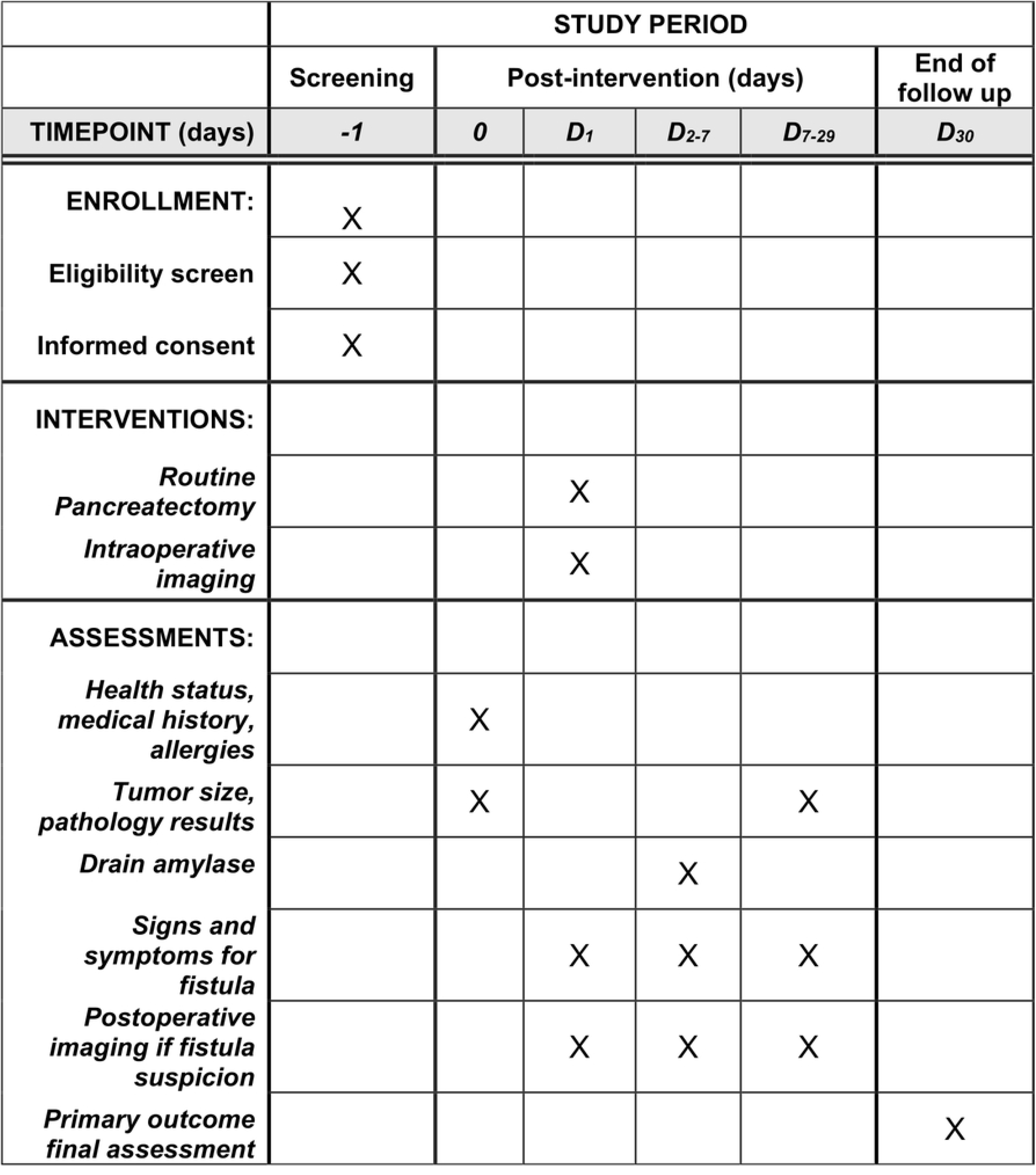
SPIRIT Schedule of Enrollment Interventions and Assessments

### Outcomes

#### Intraoperative

During the surgery, a 90-120-second video will be captured to analyze ICG perfusion metrics. Because no current gold-standard metric exists for this imaging technique in the pancreas, several metrics will be analyzed. Quantitative ICG metrics will include absolute and relative ROI scores within the Cinevaq (Stryker, USA) software, T0 (the time from ICG injection until the first fluorescent signal), TMax (time from the first ICG signal until maximum uptake is achieved), as well as ingress and egress rates for the contrast [19,20].

#### Postoperative

After the surgery, the participant would enter the study’s follow-up period. The main outcome will be the development or absence of postoperative pancreatic fistula, as defined by the most updated definition of the International Study Group in Pancreatic Surgery (ISGPS) [21].

Following this, we will collect surgical drain amylase levels in the first three days after surgery, clinical conditions of the participant, imaging results, and persistent drainage from the drain, among other factors delineated in the ISGPS. Figure 2 depicts a flowchart for individual participants in the trial.

**Figure 2.**
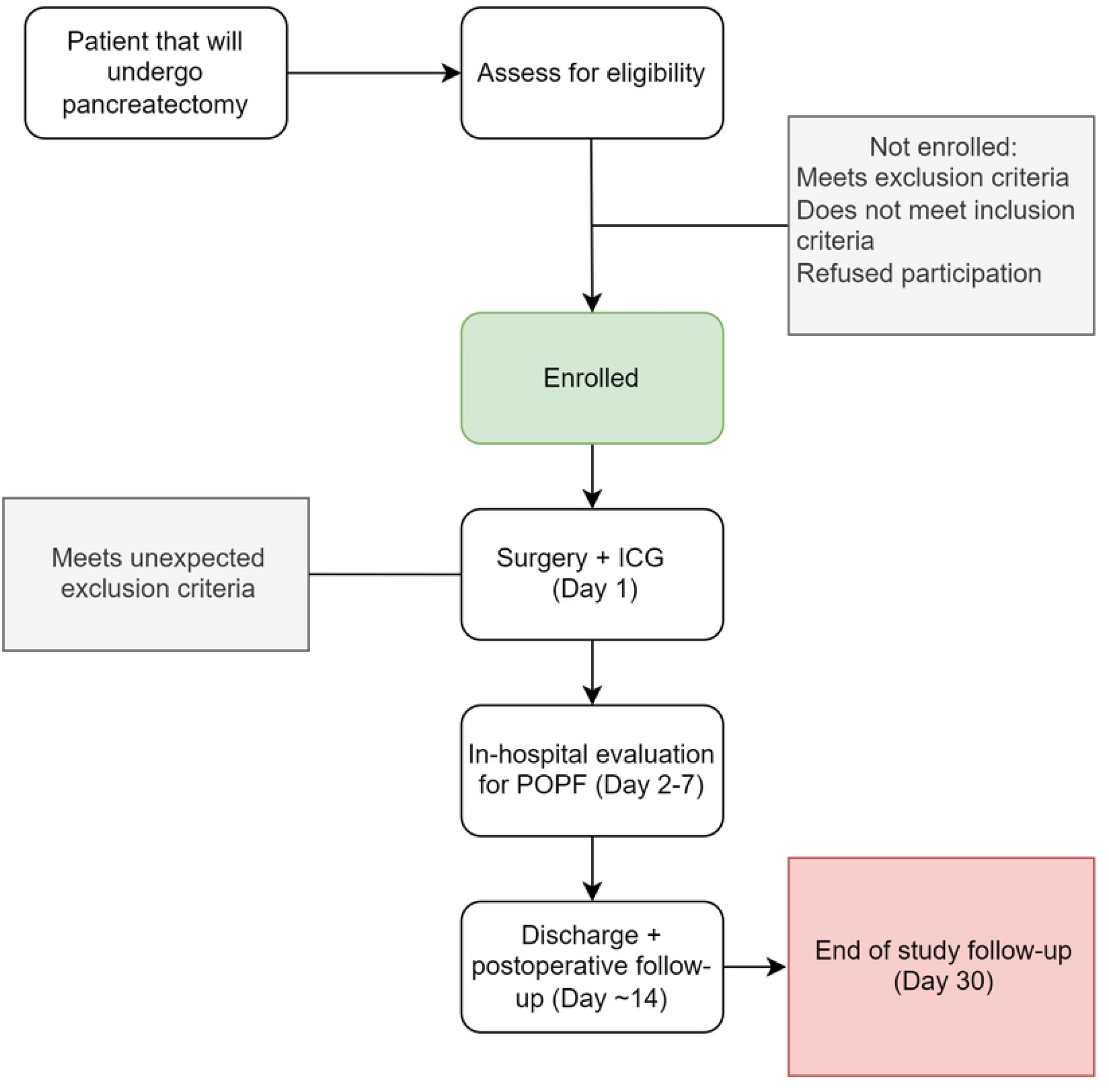
Trial Flowchart. Flowchart for participants in the VIPR-1 trial.

Descriptive statistics will be used to compare patients that developed postoperative fistula. ICG metrics will then be compared with multivariable modeling to study the impact they might have on postoperative fistulas.

Because there are characteristics that might influence development of POPF, the following participant and case characteristics will be recorded and analyzed as covariates affecting outcomes: 1) type of pancreatectomy, 2) principal surgeon, 3) operative length, 4) blood loss in mL, 5) intraoperative blood transfusion, 6) duct size in mm, 7) gland texture, 8) intraoperative use of vasopressors, 9) race, 10) BMI, 11) age, 12) neoadjuvant therapy and 13) vascular reconstruction [22].

### Data management

Data to be collected includes baseline participant demographics relevant to our study, including sex, age, ethnicity, race, height, weight, ECOG performance status, disease stage at study entry, cancer therapy and biomarker status.

Collected data will be stored in a trial-specific database that lives in an encrypted cloud service that is institutionally compliant. Here, participant-level information will be captured without any of the 18 HIPPA identifiers; a participant key will be stored separately and securely. This study will comply with the Data and Safety Monitoring Plan established by the Knight Clinical Research Quality & Administration.

### Safety considerations

There are two relevant safety considerations for this study. The first involves safety during the imaging procedure, and the second involves securing participant data. ICG is considered a relatively inert contrast agent compared to other more commonly used imaging agents. The most frequently encountered adverse effects of ICG injection include allergic-type reactions, presenting as nausea and urticaria, with life-threatening complications such as anaphylactic shock being rare (<0.05%) [23]. Participants are made aware of the potential consequences of ICG injection at the time of screening and consent.

All adverse effects are reported within 24 hours through the Clinical Trials Management System, connected to ClinicalTrials.gov. The Knight Cancer Institute, as the trial sponsors, performs constant auditing on a participant-level basis. Participant information will be kept secure by restricting access to direct study personnel, and all images and captured participant-level data will be kept in encrypted platforms.

### Ethical considerations

The VIPR-1 trial follows guidelines set out by the Declaration of Helsinki [24]. The trial underwent review and subsequent approval from the Knight Cancer Research Institute via the Clinical Research and Review Committee on 9/2023 and then the Institutional Review Board from Oregon Health and Science University on 11/2023.

### Status and timeline

The VIPR-1 trial is registered in the ClinicalTrials government platform [25]. Participant recruitment started on 06/10/2024, and the estimated finishing time is one year after the first patient enrollment. Any updates to the protocol will be informed in the trial registry platform and the protocol itself. After the trial is completed, we intend to publish our results in a scientific journal.

## Discussion

This is the first prospective clinical trial in the United States to evaluate pancreatic remnant perfusion using ICG to predict fistulas and leaks. A recently published systematic review compiled all reports of the use of pancreatic remnant perfusion intraoperatively. A total of 5 studies were found, and only two were prospective [26]. The first prospective trial used ICG for postoperative acute pancreatitis rather than POPF as a primary outcome measure, where adequate statistical power for POPF was not mentioned [14]. The other prospective trial looked at subjective descriptions of adequate blood flow to the pancreatic remnant to characterize hypoperfusion and guide margins [27]. Thus, further highlighting the need for prospective intraoperative and subjective evaluation of perfusion. While studies have reported that hypoperfusion detected by ICG is correlated with POPF, currently, no cutoff for considering adequate or inadequate perfusion exists. After operative measurements are performed, we intend to utilize the Cinevaq software (Stryker, USA) to measure the perfusion difference with reference to gastrointestinal tissue and within the pancreas. If a correlation between perfusion and leaks is detected, we will perform statistical analysis at different thresholds for poor perfusion to identify the best cutoff delta in perfusion to predict POPF. A recent study demonstrated that cutoff values for hypoperfusion in breast tissue do not translate between different imaging systems. Specifically, a 33% relative perfusion measurement has been validated in breast tissue, specifically in the SPY-Elite® platform. However, on the newer SPY-PHI platform, the same cutoff value did not correlate with hypoperfusion [28]. While studies like this are not available in pancreatic tissue, it is possible that a specific cutoff would need to be validated for each type of device model in pancreatic perfusion dynamics.

If the results are positive, a phase III study can be designed to evaluate whether ICG metrics can guide surgical margins to optimize perfusion of the pancreatic remnant and decrease POPF. The level of the transection of the neck of the pancreas has been previously characterized as important for POPF risk [29]. Capturing patient preoperative and intraoperative variables is crucial to adjust for possible confounders in perfusion observations and the development of POPF. All patients undergoing open pancreatectomy for any diagnosis will be screened for this study. Broad inclusion and exclusion criteria will be implemented to ensure a diverse group of patients and reach the sample size needed.

In the past decade, minimally invasive pancreatectomy has gained major interest, as evidenced by numerous studies examining ways to reduce postoperative complications. Current data are inconclusive regarding the impact of surgical technique (open versus minimally invasive) on POPF rates, with some studies suggesting benefits. In contrast, others indicate an increased risk of this outcome with minimally invasive surgery [16,30]. A recent proof of concept prospective study investigated the predictive potential of ICG in determining suture tension-induced hypoperfusion in pancreatic reconstruction. In this study, hypoperfusion to the pancreas by ICG was more frequently encountered in those patients who developed fistulas [31]. This study, however, was performed only in robotic cases, and all measurements were taken after the surgeon placed sutures for the anastomosis. We intend to measure perfusion before the pancreas is sutured to the jejunum to measure the complete surface of the remnant to be anastomosed.

Pancreatectomies are among the most challenging and lengthy procedures in gastrointestinal surgery. Because of this, the trial’s impact on time added to the case should be minimal. Increases in operative time elevate costs and staff utilization and are associated with surgical complications in pancreatic surgery [32]. To address this, the operative time will be compared to control cases that did not undergo ICG measurements. This comparison will determine the impact of ICG measurements on total operative time. Studies in other gastrointestinal surgeries using ICG have shown a potential increase in operative time by 10-15 minutes, but this increase has not been statistically significant [33]. We currently do not know the impact of ICG in open pancreatectomies. ICG is now standard in many gastrointestinal procedures but not for the pancreas. We intend to record and subsequently report the effect of ICG on operative time for pancreatectomies.

The main limitation of this study is that our findings might not be generalizable to other institutions because of inherent differences in practice standards, surgeon experience, and imaging devices used.

In conclusion, POPF is highly prevalent in pancreatectomy. Current predictive measures to detect POPF intraoperatively have not been successful in decreasing complication rates. ICG perfusion of the pancreatic edge could potentially allow surgeons to predict POPF. We aim to investigate this further with the VIPR-1 trial.

## Data Availability

No datasets were generated or analysed during the current study. All relevant data from this study will be made available upon study completion.

## Authors Contributions - CRediT author statement

GSG: Writing-Original Draft Preparation, Methodology. AW: Writing-Original draft preparation, FGR: Writing-Reviewing and Editing, SCM: Visualization, Writing-Reviewing and Editing, BCS: Visualization, Writing-Reviewing and Editing, PJW: Conceptualization, Methodology, Writing – Original draft, Funding Acquisition, Project Administration.

## Acknowledgments

None.

## Supporting information

S1 Fig. **SPIRIT Checklist**.

## References

1. Cancer Facts & Figures 2023 [Internet]. [cited 2024 Apr 9]. Available from: https://www.cancer.org/research/cancer-facts-statistics/all-cancer-facts-figures/2023-cancer-facts-figures.html

2. Strobel O, Lorenz P, Hinz U, Gaida M, König AK, Hank T, et al. Actual Five-year Survival After Upfront Resection for Pancreatic Ductal Adenocarcinoma: Who Beats the Odds? Ann Surg. 2022 May 1;275(5):962–71.

3. Malgras B, Dokmak S, Aussilhou B, Pocard M, Sauvanet A. Management of postoperative pancreatic fistula after pancreaticoduodenectomy. J Visc Surg. 2023 Feb 1;160(1):39–51.

4. Torres OJM, Moraes-Junior JMA, Fernandes EDSM, Hackert T. Surgical Management of Postoperative Grade C Pancreatic Fistula following Pancreatoduodenectomy. Visc Med. 2022;38(4):233–42.

5. Dhayat SA, Tamim ANJ, Jacob M, Ebeling G, Kerschke L, Kabar I, et al. Postoperative pancreatic fistula affects recurrence-free survival of pancreatic cancer patients. Chen RJ, editor. PLOS ONE. 2021 Jun 4;16(6):e0252727.

6. Bonaroti JW, Zenati MS, Al-abbas AI, Rieser CJ, Zureikat AH, Hogg ME, et al. Impact of postoperative pancreatic fistula on long-term oncologic outcomes after pancreatic resection. HPB. 2021 Aug;23(8):1269–76.

7. Callery MP, Pratt WB, Kent TS, Chaikof EL, Vollmer CM. A Prospectively Validated Clinical Risk Score Accurately Predicts Pancreatic Fistula after Pancreatoduodenectomy. J Am Coll Surg. 2013 Jan;216(1):1–14.

8. Nahm CB, Brown KM, Townend PJ, Colvin E, Howell VM, Gill AJ, et al. Acinar cell density at the pancreatic resection margin is associated with post-pancreatectomy pancreatitis and the development of postoperative pancreatic fistula. HPB. 2018 May;20(5):432–40.

9. Nahm CB, Connor SJ, Samra JS, Mittal A. Postoperative pancreatic fistula: a review of traditional and emerging concepts. Clin Exp Gastroenterol. 2018 Mar 15;11:105–18.

10. De Muynck LDAN, White KP, Alseidi A, Bannone E, Boni L, Bouvet M, et al. Consensus Statement on the Use of Near-Infrared Fluorescence Imaging during Pancreatic Cancer Surgery Based on a Delphi Study: Surgeons’ Perspectives on Current Use and Future Recommendations. Cancers. 2023 Jan 20;15(3):652.

11. Newton AD, Predina JD, Shin MH, Frenzel-Sulyok LG, Vollmer CM, Drebin JA, et al. Intraoperative Near-infrared Imaging Can Identify Neoplasms and Aid in Real-time Margin Assessment During Pancreatic Resection. Ann Surg. 2019 Jul;270(1):12–20.

12. Asbun D, Kunzler F, Marin R, Asbun HJ. Pancreatic fluorescence using continuous indocyanine green infusion. J Surg Oncol. 2022 Dec;126(7):1215–8.

13. Rho SY, Kim SH, Kang CM, Lee WJ. Is ICG-enhanced image able to help predicting pancreatic fistula in laparoscopic pancreaticoduodenectomy? Minim Invasive Ther Allied Technol MITAT Off J Soc Minim Invasive Ther. 2019 Feb;28(1):29–32.

14. Doussot A, Decrock M, Calame P, Georges P, Turco C, Lakkis Z, et al. Fluorescence-based pancreas stump perfusion is associated with postoperative acute pancreatitis after pancreatoduodenectomy a prospective cohort study. Pancreatol Off J Int Assoc Pancreatol IAP Al. 2021 May 18;S1424-3903(21)00161-7.

15. Hope-Ross M, Yannuzzi LA, Gragoudas ES, Guyer DR, Slakter JS, Sorenson JA, et al. Adverse Reactions due to Indocyanine Green. Ophthalmology. 1994 Mar;101(3):529–33.

16. Panni RZ, Panni UY, Liu J, Williams GA, Fields RC, Sanford DE, et al. Re-defining a high volume center for pancreaticoduodenectomy. HPB. 2021 May;23(5):733–8.

17. El Amrani M, Clément G, Lenne X, Laueriere C, Turpin A, Theis D, et al. Should all pancreatic surgery be centralized regardless of patients’ comorbidity? HPB. 2020 Jul;22(7):1057–66.

18. Chan AW, Tetzlaff JM, Altman DG, Laupacis A, Gøtzsche PC, Krleža-Jerić K, et al. SPIRIT 2013 Statement: Defining Standard Protocol Items for Clinical Trials. Ann Intern Med. 2013 Feb 5;158(3):200.

19. Oppermann C, Dohrn N, Pardes HY, Klein MF, Eriksen T, Gögenur I. Real time organ hypoperfusion detection using Indocyanine Green in a piglet model. Surg Endosc [Internet]. 2024 Jun 13 [cited 2024 Jul 15]; Available from: https://link.springer.com/10.1007/s00464-024-10938-0

20. Iwamoto H, Matsuda K, Hayami S, Tamura K, Mitani Y, Mizumoto Y, et al. Quantitative Indocyanine Green Fluorescence Imaging Used to Predict Anastomotic Leakage Focused on Rectal Stump During Laparoscopic Anterior Resection. J Laparoendosc Adv Surg Tech. 2020 May 1;30(5):542–6.

21. Bassi C, Marchegiani G, Dervenis C, Sarr M, Abu Hilal M, Adham M, et al. The 2016 update of the International Study Group (ISGPS) definition and grading of postoperative pancreatic fistula: 11 Years After. Surgery. 2017 Mar;161(3):584–91.

22. Schuh F, Mihaljevic AL, Probst P, Trudeau MT, Müller PC, Marchegiani G, et al. A Simple Classification of Pancreatic Duct Size and Texture Predicts Postoperative Pancreatic Fistula: A classification of the International Study Group of Pancreatic Surgery. Ann Surg. 2023 Mar;277(3):e597.

23. Obana A, Miki T, Hayashi K, Takeda M, Kawamura A, Mutoh T, et al. Survey of Complications of Indocyanine Green Angiography in Japan. Am J Ophthalmol. 1994 Dec;118(6):749–53.

24. World Medical Association Declaration of Helsinki: Ethical Principles for Medical Research Involving Human Subjects. JAMA. 2013 Nov 27;310(20):2191.

25. Worth PJ. Assessing the Role of Intraoperative Indocyanine Green Perfusion of the Transected Pancreas in Predicting Postoperative Pancreatic Leaks [Internet]. clinicaltrials.gov; 2024 Mar [cited 2023 Dec 31]. Report No.: NCT06084013. Available from: https://clinicaltrials.gov/study/NCT06084013

26. Robertson FP, Spiers HVM, Lim WB, Loveday B, Roberts K, Pandanaboyana S. Intraoperative pancreas stump perfusion assessment during pancreaticoduodenectomy: A systematic scoping review. World J Gastrointest Surg. 2023 Aug 27;15(8):1799–807.

27. Strasberg SM, Drebin JA, Mokadam NA, Green DW, Jones KL, Ehlers JP, et al. Prospective Trial of a Blood Supply-Based Technique of Pancreaticojejunostomy: Effect on Anastomotic Failure in the Whipple Procedure1. J Am Coll Surg. 2002 Jun;194(6):746–58.

28. Lauritzen E, Bredgaard R, Bonde C, Jensen LT, Damsgaard TE. An observational study comparing the SPY-Elite® vs. the SPY-PHI QP system in breast reconstructive surgery. Ann Breast Surg. 2023 Jun;7:12–12.

29. Bardol T, Delicque J, Hermida M, Herrero A, Guiu B, Fabre JM, et al. Neck transection level and postoperative pancreatic fistula after pancreaticoduodenectomy: A retrospective cohort study of 195 patients. Int J Surg. 2020 Oct;82:43–50.

30. Uijterwijk BA, Kasai M, Lemmers DHL, Chinnusamy P, Van Hilst J, Ielpo B, et al. The clinical implication of minimally invasive versus open pancreatoduodenectomy for non-pancreatic periampullary cancer: a systematic review and individual patient data meta-analysis. Langenbecks Arch Surg. 2023 Aug 15;408(1):311.

31. Chen JW, Lof S, Zwart MJW, Busch OR, Daams F, Festen S, et al. Intraoperative Fluorescence Imaging During Robotic Pancreatoduodenectomy to Detect Suture-Induced Hypoperfusion of the Pancreatic Stump as a Predictor of Postoperative Pancreatic Fistula (FLUOPAN): Prospective Proof-of-concept Study. Ann Surg Open. 2023 Dec;4(4):e354.

32. Williams MD, Bhama AR, Naffouje S, Kamarajah SK, Becerra AZ, Zhang Y, et al. Effect of Operative Time on Outcomes of Minimally Invasive Versus Open Pancreatoduodenectomy. J Gastrointest Surg. 2023 Jan;27(1):93–104.

33. Cassinotti E, Al-Taher M, Antoniou SA, Arezzo A, Baldari L, Boni L, et al. European Association for Endoscopic Surgery (EAES) consensus on Indocyanine Green (ICG) fluorescence-guided surgery. Surg Endosc. 2023 Mar;37(3):1629–48.

